# Diagnostic accuracy of Impedance Spectroscopy versus Digital Rectal Examination for Obstetric Anal Sphincter Injuries: a postpartum post-hoc analysis

**DOI:** 10.1101/2024.09.18.24313868

**Authors:** Stefano Salvatore, Katarzyna Borycka, Alessandro Ruffolo, Marcel Młyńczak, Maciej Rosoł, Kacper Korzeniewski, Piotr Iwanowski, Antonino Spinelli, Renaud DeTayrac, Carlo Ratto, Stavros Athanasiou, Diaa Rizk, Andrea Stuart, Jan Baekelandt, Małgorzata Uchman-Musielak, Hynek Heřman, Petr Janku, Peter Rosenblat, Mariusz Grzesiak, Adam Dziki, Ruwan Fernando

**Author notes:** Corresponding author: Katarzyna Borycka, MD, PhD, Postal address: Department of General Surgery, Medicover Hospital, 5 Rzeczypospolitej Street, Warsaw, Poland.

## Abstract

**Background:** Accurate diagnosis of obstetric anal sphincter injuries (OASIs) is critical for timely repair and prevention of long-term morbidity, yet digital rectal examination (DRE) remains insufficiently sensitive.

**Methods:** In this post-hoc analysis of a prospective multicentre diagnostic study (NCT04903977), we compared the diagnostic performance of DRE and machine learning-assisted impedance spectroscopy with three-dimensional endoanal ultrasound (EAUS) as the reference. A total of 152 women who delivered vaginally were assessed up to 8 weeks postpartum.

**Results:** DRE demonstrated a sensitivity of 44.3%, specificity of 83.5%, and overall accuracy of 67.8%. Impedance spectroscopy achieved 90.6% sensitivity, 84.6% specificity, and 87.0% accuracy against EAUS.

**Conclusions:** DRE frequently fails to detect OASIs postpartum. A minimally invasive, operator-independent impedance-based method significantly improves diagnostic sensitivity and warrants further evaluation for use immediately after delivery to enable timely primary repair.

## Introduction

Obstetric anal sphincter injuries (OASIs) represent a major clinical challenge in maternal care [1–10]. Despite being widely underdiagnosed, they are strongly associated with long-term complications, including faecal incontinence, pain, and pelvic floor dysfunction [1,8] The reported incidence of OASI ranges widely, from 0.1% to 6% in routine clinical records [4,5], yet population-level imaging studies suggest a true prevalence exceeding 25% [11]. Early detection and appropriate repair of OASI ideally within 8-12 hours post-delivery is critical to achieving optimal outcomes and avoiding delayed surgical intervention[12–14].

Digital rectal examination (DRE), although widely accessible, and recommended by guidelines [12–24], shows poor sensitivity and high operator dependency [2,25]. Endoanal ultrasound (EAUS), considered the reference standard for OASI detection, is limited in routine delivery settings due to its cost, availability, and technical challenges when used immediately postpartum. In particular, studies have shown that edema, hematoma, and gas artifacts compromise the reliability of EAUS in the hours following delivery [26]

Alternative imaging approaches, such as transperineal or introital ultrasound, are easier to apply and use standard obstetric equipment [27]. However, they lack precision in grading sphincter defects and are not currently endorsed by international guidelines [17, 28]. This creates a critical diagnostic gap during the window when timely repair is most effective.

To address this gap, we evaluated a machine learning-based impedance spectroscopy system designed to detect structural changes in the anal sphincter complex. While previous pilot studies have demonstrated the feasibility of this technique [29–31], the current post-hoc analysis from the pivotal study [32] assesses its diagnostic performance in a controlled early postpartum setting, using 3D-EAUS as the reference standard.

Our objective was to compare the sensitivity, specificity, and diagnostic accuracy of impedance spectroscopy and DRE, focusing on their ability to identify OASI in the postpartum period when EAUS reliability is optimal. These results may inform future use of impedance-based diagnostics as a complementary tool to clinical examination in postpartum care.

## Materials and Methods

### Study Design and Population

This post-hoc analysis is based on data from a prospective, comparative, multicentre diagnostic study [32] conducted between May 2021 and December 2022 at five European centres. The study was designed to validate the diagnostic performance and safety of a machine learning-assisted impedance spectroscopy system for detecting OASI, using three-dimensional endoanal ultrasound (EAUS) as the reference standard (ClinicalTrials.gov identifier: NCT04903977).

Women aged 18-49 who delivered vaginally at ≥34 weeks gestation were eligible for inclusion. Both primiparous and multiparous women were enrolled. Exclusion criteria included: (1) pre-existing faecal incontinence unrelated to OASI; (2) prior perineal or anal surgery; (3) active inflammatory bowel disease; (4) cardiac devices or arrhythmias; and (5) any acute or chronic conditions that would interfere with study procedures.

The study adhered to ISO 14155:2020 for good clinical practice in medical device trials and followed the principles of the Declaration of Helsinki. Ethical approval was obtained at each centre (see Declarations)*, and written informed consent was obtained from all participants.

### Study Procedures

Participants were recruited within 8 weeks postpartum, allowing assessments to be performed after the resolution of acute delivery-related oedema and bleeding. The study included three visits:

-Visit 1: Clinical history, physical examination, 12-lead ECG, Wexner incontinence score, and 3D-EAUS.

-Visit 2: Within 7 days of Visit 1, participants underwent impedance spectroscopy and digital rectal examination (DRE).

-Visit 3: Follow-up safety assessment (not included in this analysis).

### Reference and Index Tests

Three-dimensional endoanal ultrasound (EAUS) served as the reference standard. Scans were performed using a 360° 3D endoprobe by experienced colorectal specialists. EAUS results were evaluated using three validated scales: the OASIS classification (grades 3a-4), the Norderval score (quantitative sphincter defect), and a modified Starck classification (anatomical mapping of internal and external sphincter injury). Results were finalized and electronically locked before impedance test was performed.

DRE was conducted bi-digitally following international guidelines [12], with the result recorded as either “OASI suspected” or “OASI not suspected.” All examiners had prior experience in perineal trauma assessment.

Impedance spectroscopy was performed using a single-use endoanal probe connected to a handheld spectrometer (ONIRY system). Measurements lasted under one minute in lithotomy position. The embedded machine learning algorithm returned a binary output (“PASS or REFER”) without operator interpretation or external processing.

### Blinding

Clinicians performing EAUS were blinded to the results of both index tests. DRE examinations were conducted by trained investigators who were not involved in EAUS assessment. The impedance spectroscopy system provided fully automated classification based on internal algorithms, eliminating the need for blinding. Operator-independent processing ensures reproducibility and minimizes bias in impedance-based results.

### Device Description and ML Model

The ONIRY system includes an impedance spectrometer and a single-use endoanal probe, which records tissue impedance in a supine, gynaecological position over approximately one minute. Impedance data are analysed using an embedded neural network trained to detect tissue patterns associated with OASI. The device provides a binary output: “PASS” (no OASI) or “REFER” (suspected OASI), along with probable injury localization. All ML processing occurs within the device; no external software is required.

The model was validated using 10 iterations of 10-fold cross-validation on impedance data, classified against the EAUS reference.

### Independent Factors Assessed

To understand factors affecting impedance readings, potential confounders such as residual fluids, blood, and inflammation (monitored via calprotectin levels) were independently analysed. This approach ensures the robustness of the impedance data and supports future refinement of ONIRY performance in early postpartum settings. Follow-up studies focused exclusively on day 1 postpartum use are ongoing.

### Post-Hoc Grouping

Participants were reclassified into two groups based on EAUS findings:

- Group I (No OASI): Including intact perineum, first-or second-degree perineal tears

- Group II (OASI present): Third-or fourth-degree sphincter injuries Statistical Analysis

Diagnostic performance was calculated for each index test using EAUS as the reference standard. Primary outcomes were sensitivity, specificity, and accuracy. They were categorized as Success (True Positive, TP or True Negative, TN) or Failure (False Positive, FP, or False Negative, FN). Secondary analysis included performance by OASI grade (3a-4) and area under the receiver operating characteristic (ROC) curve.

Cross-validation metrics for the machine learning model were reported as mean ± standard deviation across 10 iterations.

## Results

A total of 152 women were included in the final analysis. Baseline demographic and clinical characteristics are presented in Table 1.

**Table 1.** Characteristics of the study population.

|  | Group I | Group II | Total |
| --- | --- | --- | --- |
|  | (N=91) | (N=61) | (N=152) |
| Age (years) |  |  |  |
| Mean ± SD | 31.7 ± 4.6 | 30.7 ± 4.6 | 31.3 ± 4.6 |
| Range (Min/Max) | 22 (18/40) | 20 (22/42) | 24 (18/42) |
| Age categorized (n, %) |  |  |  |
| <26 years | 9 (9.9) | 7 (11.5) | 16 (10.5) |
| 26<35 years | 53 (58.2) | 41 (67.2) | 94 (61.8) |
| ≥35 years | 29 (31.9) | 13 (21.3) | 42 (27.6) |
| Weight (kg) |  |  |  |
| Mean ± SD | 71.7 ± 11.8 | 68.1 ± 9.6 | 70.2 ± 11.0 |
| Range (Min/Max) | 56 (49/105) | 60 (48/108) | 60 (48/108) |
| BMI (kg/m<sup>2</sup>) |  |  |  |
| Mean ± SD | 25.4 ± 3.6 | 24.3 ± 3.4 | 25.0 ± 3.6 |
| Range (Min/Max) | 18.9 (17.4/36.3) | 15.6 (17.7/33.3) | 18.9 (17.4/36.3) |
| Number of pregnancies (including the index one) (n) |  |  |  |
| Median | 2 | 1 | 1 |
| Range (Min/Max) | 5 (1/6) | 3 (1/4) | 5 (1/6) |
| Primipara/Multipara | 49/42 | 46/15 | 95/57 |
| Risk factors for Obstetric Anal Sphincter Injury from the index delivery (n, %) |  |  |  |
| Prolonged second phase of delivery | 6 (6.6) | 10 (16.4) | 16 (10.5) |
| Fetal shoulder dystocia | 1 (1.1) | 0 (0.0) | 1 (0.7) |
| Birth weight of the neonate >4kg | 9 (9.9) | 6 (9.8) | 15 (9.9) |
| Induction of delivery with oxytocin | 15 (16.5) | 9 (14.8) | 24 (15.8) |
| Head circumference of the neonate ≥34 cm | 56 (61.53) | 49 (80.3) | 105 (69.1) |
| Time between the index delivery and ONIRY examination (days) |  |  |  |
| Median | 4 | 28 | 14 |
| Range (Min/Max) | 57 (0/57) | 55 (1/56) | 57 (0/57) |
| Time between the index delivery and EAUS examination (days) |  |  |  |
| Median | 4 | 28 | 14 |
| Range (Min/Max) | 57 (0/57) | 55 (1/56) | 57 (0/57) |

Group reallocation based on EAUS revealed that 61 participants had confirmed OASI (Group II), while 91 participants had no OASI (Group I).

The diagnostic accuracy, sensitivity, and specificity of the DRE and ONIRY systems compared to EAUS as the reference standard were evaluated. The performance of DRE in identifying OASIs indicated an accuracy of 67.8%, reflecting the proportion of total diagnoses (both positive and negative) that were correctly identified. Sensitivity, which measured the ability of DRE to correctly identify those with the OASI presence (true positive rate), was relatively low at 44.3%. On the other hand, specificity, measuring the ability to correctly identify those without the condition (true negative rate), was higher at 83.5%.

The machine learning-assisted impedance spectroscopy was evaluated by comparing the trained ML model’s output with EAUS results. The system achieved an accuracy of 87.0% ± 0.5%, which represents a significant enhancement in the overall ability to diagnose OASIs correctly. The sensitivity of the ONIRY system was 90.6% ± 2.0%, indicating its strong capability to identify true positive cases of OASIs effectively. Specificity was robust at 84.6% ± 1.9%, suggesting that the ONIRY system maintains a high level of accuracy in confirming the absence of injuries, similar to DRE.

The ONIRY performance metrics for each iteration of 10-fold cross-validation and the overall statistical analysis are given in Table 2.

**Table 2.** Performance metrics of the ONIRY device in the assessment relative to 3-D Endoanal Ultrasound and OASIS classification (each row shows the statistics for a single 10-fold cross-validation each performed with different random seed).

| Seed | Accuracy | Sensitivity | Specificity |
| --- | --- | --- | --- |
| 1 | 87.2% | 90.9% | 84.6% |
| 2 | 86.6% | 92.6% | 82.3% |
| 3 | 87.9% | 86.7% | 88.6% |
| 4 | 86.9% | 87.7% | 86.4% |
| 5 | 87.6% | 92.7% | 84.2% |
| 6 | 87.6% | 89.4% | 86.2% |
| 7 | 86.9% | 91.9% | 83.5% |
| 8 | 86.2% | 91.0% | 82.9% |
| 9 | 86.9% | 91.9% | 83.5% |
| 10 | 86.6% | 91.2% | 83.6% |
| Mean ±SD | 87.0% ± 0.5% | 90.6% ± 2.0% | 84.6% ± 1.9% |

The accuracy of the two compared methods in detecting OASI in each OASIS classification grades 3a, 3b, 3c, and 4 is shown in Table 3.

**Table 3.** Performance metrics of DRE and ONIRY device in the assessment in reference to EAUS under OASIS classification.

| Grade | N | Accuracy of DRE | Accuracy of impedance spectroscopy |
| --- | --- | --- | --- |
| Grade 3a | 25 | 24.0% | 81.2% |
| Grade 3b | 26 | 53.8% | 90.8% |
| Grade 3c | 9 | 77.8% | 95.6% |
| Grade 4 | 1 | 0.0% | 60% |

The comparison of receiver operating characteristic (ROC) curves for ONIRY and DRE, along with their respective area under the curve (AUC) values, is presented in Figure 1.

**Figure 1.**
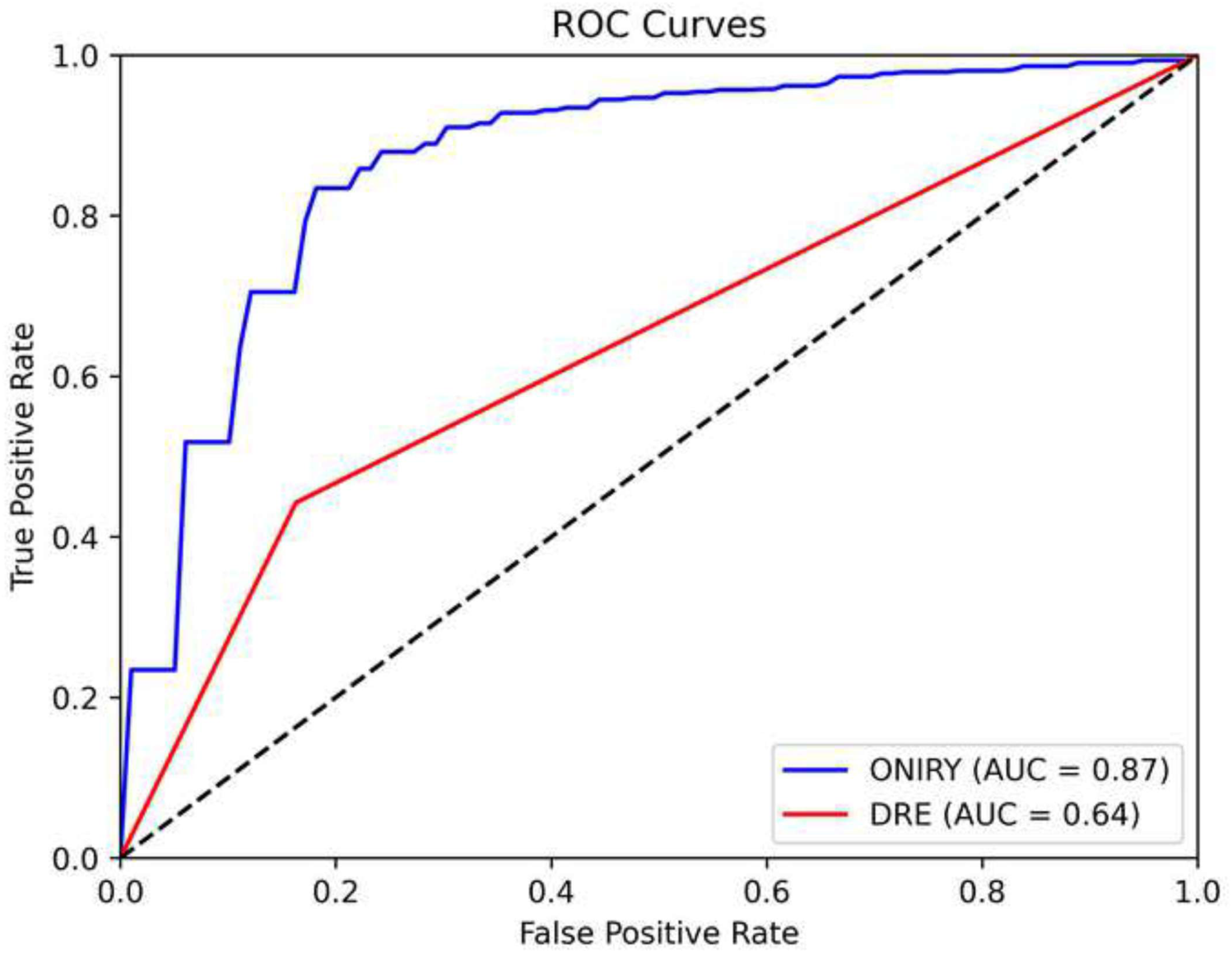
ROC curves for the ONIRY system and DRE are presented as blue and red lines, respectively, with the identity line depicted as a black dashed line.

## Discussion

This post-hoc analysis compared the diagnostic performance of DRE and machine learning-assisted impedance spectroscopy in detecting OASIs, using three-dimensional endoanal ultrasound (EAUS) as the reference standard. The results demonstrate a significant difference in sensitivity between the two methods: DRE correctly identified only 44.3% of confirmed OASIs, whereas impedance spectroscopy achieved a sensitivity of 90.6%, with overall diagnostic accuracy exceeding 87%.

DRE remains a first-line tool in postpartum care due to its accessibility, yet its performance is highly dependent on examiner training and experience. DRE was more effective in confirming the absence of injury than detecting its presence and had the poorest accuracy in assessing the extent of external sphincter injury (24% and 53.8% in OASIS grades 3a and 3b, respectively). Our results reinforce prior studies [25,33] showing that DRE frequently fails to detect external sphincter injuries, especially grade 3a and 3b tears, which often present subtly on palpation. This is particularly concerning given that accurate differentiation between second- and third-degree tears is essential for determining the need for surgical repair. And this is what is most challenging for clinicians in obstetric practice: how to differentiate between perineal injury grade 2 which can be addressed by a midwife, and grade 3, when the external sphincter is injured, and an experienced obstetrician (or colorectal surgeon) with expertise in OASI is required for repair.

Impedance spectroscopy, by contrast, provides objective, operator-independent classification based on biophysical tissue properties. In this study, it was able to detect OASI with high sensitivity across all injury grades, including permance was validated using cross-validated machine learning on study-derived data, enhancing reproducibility. While EAUS remains the imaging reference standard, its practical limitations especially immediately after delivery due to oedema, bleeding, and anatomical distortion are well-documented [26], and limit its availability in real-world delivery settings.

Importantly, this study did not evaluate the performance of impedance spectroscopy in the immediate postpartum period. Instead, the analysis was deliberately conducted during the early postpartum window (up to 8 weeks post-delivery) to optimize EAUS reliability and enable robust training of the machine learning model. This design enhances internal validity but limits generalizability to the delivery room setting.

The clinical consequences of undiagnosed or misclassified OASIs are substantial. Delayed recognition often leads to incomplete or suboptimal repair, resulting in persistent sphincter defects and long-term morbidity. Meta-analyses by Okeahialam et al. [34] report rates of anal incontinence exceeding 18% following OASI, with higher rates among more severe tear grades (15.6% for 3a, 18.3% for 3b, 20.6% for 3c, and 28.4% for 4th-degree tears). Taithongchai et al. [35]and Roper et al.)[36] have shown that missed or underdiagnosed OASIs are associated with worse continence outcomes and higher need for secondary colorectal procedures. In contrast, structured repair immediately after delivery is associated with favourable continence (7% pof fecal incontinence, with an additive 24% rate of mild gas incontinence symptoms) and quality-of-life outcomes [37,38].

Cost-effectiveness analyses also favor early diagnosis demonstrated that primary repair leads to both improved quality-adjusted life years and lower long-term healthcare costs compared with delayed sphincteroplasty [39]. These findings support efforts to strengthen postpartum OASI detection, particularly during the therapeutic window in which surgical repair can be most effective.

This study’s strengths include its multicentre design, use of a high-quality imaging reference standard, and blinded EAUS interpretation. However, several limitations must be acknowledged. First, the diagnostic accuracy of DRE may be artificially high in this study due to examiner expertise, which may exceed that of typical maternity settings. Second, the impedance spectroscopy model was trained and validated on the same study population; while cross-validation mitigates overfitting, external validation is needed. Finally, the study does not address the diagnostic performance of the impedance system in the immediate postpartum window a critical time for enabling primary repair.

Future research should focus on validating impedance spectroscopy in real-time delivery settings, ideally in prospective cohorts where early intervention decisions depend on rapid and reliable detection. Additionally, comparative studies with transperineal ultrasound may help define the role of impedance-based methods within the broader spectrum of postpartum diagnostic tools.

### Conclusion

Digital rectal examination identified only 44% of obstetric anal sphincter injuries in the early postpartum period, while impedance spectroscopy achieved over 90% sensitivity and 87% accuracy compared with 3D endoanal ultrasound. As a minimally invasive, operator-independent method, it may offer critical diagnostic support when injury cannot be confidently excluded on clinical examination. These findings justify further investigation of impedance spectroscopy in the immediate postpartum setting, where timely detection is essential to enable primary repair and prevent long-term complications.

## Data Availability

The data underlying this paper are not publicly available as they constitute the intellectual property of the company.

## Acknowledgments

The authors would like to thank all the other investigators and co-investigators involved in the clinical trial from centres in the Czech Republic (Brno and Prague), Slovakia (Kosice), Poland (Warsaw), and Spain (Leon) for their invaluable contributions to patient recruitment, clinical trials conduct, and assistance with results.

## Disclosure of interests

A.S. is an independent Ethicon, Takeda, Pfizer, and Sofar consultant.

M.U.M, H.H. and P.J. received remuneration as study investigators.

S.S., R.DT., A.S., H.H., C.R., S.A., D.R., A.St., J.B., M.G., A.D., and R.F. are independent consultants of OASIS Diagnostics’ Scientific Advisory Board.

K.B. is an independent consultant, and a Takeda trainer; a founder and management board member at OASIS Diagnostics, the author of the related patent.

## Contribution to Authorship

Conceptualization: S.S., R.F. K.B., M.M., P.I.

Data curation: S.S., M.M., M.R., K.K., P.I.

Formal analysis: M.M., M.R., K.K.

Funding acquisition: K.B., M.M.

Investigation: M.U.M., H.H., P.J.

Methodology: S.S., R.DT., A.S., H.H., C.R., S.A., D.R., A.St., J.B., R.Fr., M.G., A.D., and R.F.

Project administration: K.B., M.M. Resources: H.H., P.J.

Software: M.R., K.K.

Supervision: R.DT., A.S., H.H., C.R., S.A., D.R., A.St., J.B., R.Fr., M.G., A.D., and R.F.

Validation: M.M., P.I. Visualization: M.R., K.K.

Writing the original draft: S.S., K.B., A.R., A.S. M.M., M.R., K.K. Editing the final draft: A.St., D.R., M.U.M., A.R.

## *Details of Ethics Approval

The study received approval from the ethics committees of the respective study sites on the following dates: 19 March 2021 by the Ethics Committee of the Institute for Maternal and Child Care (approval no. 1/19.03.2021), 27 April 2021 by the Ethics Committee for Research with Medicines of the Health Areas of León and Bierzo (approval no. 2186), 9 June 2021 by the Ethics Committee of the University Hospital of Brno (approval no. 47/21Zdrav.), 14 October 2021 by the Ethics Committee at the Regional Medical Chamber in Warsaw (approval no. KB/1362/21), and 25 July 2022 by the Ethics Committee at AGEL Hospital Košice-Šace (approval no. ONIRY 3/2/2020).

